# Exploring Potential Barriers and Facilitators to Integrate Tuberculosis, Diabetes Mellitus, and Tobacco-Control Programs in India

**DOI:** 10.1101/2024.03.19.24304558

**Authors:** Nisha Mutalikdesai, Kajal Tonde, Kanchan Shinde, Rakesh Kumar, Surbhi Gupta, Girish Dayma, Anand Krishnan, Sanjay Juvekar, Ailiana Santosa, Nawi Ng, Rutuja Patil

## Abstract

**Background:** Integrating Tuberculosis (TB), Diabetes Mellitus (DM), and Tobacco-Control (TC) programs in India presents a promising strategy to address the triple burden of these diseases. However, limited information exists regarding the feasibility and influencing factors of such integration. This study investigates potential barriers and facilitators to integrate TB, DM, and TC programs in Ambegaon Block of Pune District in Maharashtra and Ballabgarh Block of Faridabad District in Haryana states of India.

**Methods:** Qualitative in-depth interviews were conducted between Nov 2022 to March 2023 among health workers, program managers, and stakeholders involved in program implementation (n=32). Interview guide was based on World Health Organization’s Health System Strengthening framework. Purposive sampling and snowball sampling were used to select participants. Rapid analysis of the data was done using the WHO HSS Framework.

**Results:** There were barriers and facilitators for the integration of TB-DM-TC programs observed in India. The primary challenges for integration are at the level of service delivery which is largely attributed to inadequate implementation of all three programs and negligible involvement of private practitioners in the program implementation. Moreover, inadequate referral system, insufficient infrastructure, limited resources, a shortage of trained staff, and lack of essential drugs and equipment impeded the uptake and coverage of services.

**Conclusion:** The results highlight the critical importance of addressing barriers and facilitators of implementation program in India to build a robust structure of managing the triple burden of TB, DM and tobacco control. The proposed strategies, such as sensitizing health system staff, implementing feedback and referral systems, and developing cross-program digital platforms offer a roadmap for policymakers and healthcare system managers. A multidimensional approach is vital for overcoming barriers and facilitating integration.

## Introduction

As per the World Health Organization (WHO), India accounted for more than two-thirds of the global TB cases in the year 2020. India has the highest burden of TB worldwide and the second-highest burden of Diabetes Mellitus (DM) (1). According to global burden of diseases (GBD) data for 2019, TB is still one of the top ten causes of disability-adjusted life years (DALYs), despite a recent rapid epidemiological change (2).

The triple burden of TB, DM, and tobacco addiction poses a threat to many TB-endemic countries that have not attempted to address it concurrently (3), demanding a shift from the traditional tuberculosis-centric lens to a more holistic perspective. Considering the increased global risk of infectious diseases and Non-Communicable Diseases (NCD) comorbidity, a collaborative preventative approach can be used to relieve the strain on the overcrowded health system. As a result, an integrated TB-DM-tobacco control approach may be considered effective in detecting and treating TB cases, as well as mitigating the residual burden of intersecting factors that contribute to a high annual incidence of TB morbidity and mortality.

This integration may promote a holistic approach to health measures, where the combined efforts address not only infectious diseases like TB but also chronic problems such as DM and the universal problem of tobacco use. This synergy will boost the total influence on public health. Given the prevalence of intersecting TB-DM epidemics, many countries embraced the WHO collaborative framework (4) for developing preventative TB-DM co-management programs. India also initiated TB-DM co-management operations through the National Tuberculosis Elimination Program (NTEP) but failed to create sufficient evidence discussing health system readiness and obstacles associated with the framework’s adoption (5).

The NTEP, formerly known as the Revised National TB Control Programme (RNTCP), is one of the biggest health programmes in the world. Since March 2016, India has implemented the National Programme for Prevention and Control of Cancer, Diabetes, CVD, and Stroke (NPCDCS), which was launched in 2010, now called as National Programme for Prevention & Control of Non-Communicable Diseases (NP-NCD). In line with the global framework created by WHO and the International Union Against TB and Lung Diseases in 2011, NTEP and NPCDCS created a joint framework in 2017. The National Tobacco Control Program (NTCP) was established by the Indian government in 2007-2008 as part of the 11th Five-Year Plan (FYP).

Eliminating tuberculosis by 2025 remains an ambitious yet imperative goal, considering the intricate challenges posed by socio-cultural practices and healthcare systems (5) .Our study, focusing on the integration of TB and DM programs, sheds light on critical challenges that hinder timely detection and effective management. The lack of awareness among healthcare practitioners and the general public regarding the intricate relationship between TB and DM leads to missed opportunities for early identification (6). Shared symptoms, such as fatigue, weight loss and increased susceptibility to infections further compound diagnostic complexities, delaying essential care. Our research, building upon the findings of previous studies (1,7,8), brings forth these challenges within the specific context of TB and DM integration, offering novel insights into the unique hurdles faced in achieving a comprehensive approach to healthcare.

Moreover, there exists a paucity of empirical evidence concerning nations characterized by elevated tobacco consumption and the incorporation of smoking control initiatives into the TB integration framework. This research endeavors to delve into the facilitators and barriers associated with the seamless integration of TB-DM-TC activities within the healthcare landscape of India.

## Methods

### Ethics Statement

The study received ethical approval from KEM Hospital Research Centre’s Research Ethics Committee (Reference No. KEMHRC/RVM/EC/2105) and the Institute Ethics Committee of All India Institute of Medical Sciences, New Delhi (Ref No. IEC-866/04.11.2022). All participants were contacted via email or phone and informed about purpose of the study and objectives. Those who showed willingness to participate in the study were administered a written informed consent form. We included all participants who consented to be part of the study. The data were collected from November 2022 to January 2023.

### Study Design

This qualitative study aimed to comprehensively investigate potential barriers and facilitators to integrating TB, DM, and TC programs in India from various perspectives by involving a diverse range of stakeholders. The study was conducted in two Indian districts: one each in the state of Maharashtra and another in Haryana.

Respondents were selected using purposive and snowball sampling strategies to ensure a diverse range of experiences and perspectives. A total of 32 in-depth interviews (IDI) were conducted using an in-depth interview guide to gather insights and perspectives on the feasibility of integrating the three programs. Of these, 13 participants were women and 19 were men. The study engaged participants across various stakeholder levels, including (i) national-level stakeholders (n=1, from NTEP), (ii) state-level stakeholders (N=11, 7 from NTEP, 2 from NPCDCS, and 2 from NTCP), (iii) district-level stakeholders consisting of ASHA workers, medical officers, community health officers, lab technicians, etc. (n=17, with direct involvement in the fieldwork for all the programs), (iv) researchers (n=1, from NPCDCS), and (v) non-governmental organization (NGO) representatives (n=2, from NTEP)

The data collection and analysis were guided by the World Health Organization’s (WHO) Health System Strengthening (HSS) framework (9), which outlines six building blocks for health system analysis and improvement. The in-depth interview guide comprised six sections *viz.* Service Delivery, Health Workforce, Health Information System, Financing, Governance and Leadership, and Access to Essential Medicine that investigated current service provision strategies for TB and DM care. These sections used to explore the barriers and facilitators influencing the delivery of TB/DM/TC services, and to assess the feasibility of implementing an integrated program for TB, DM and tobacco control.

The interview guide comprised open-ended questions to explore current service provision strategies for TB and DM care, identified barriers and facilitators affecting the delivery of TB/DM/TC services, and assessed the feasibility of rendering integrated programs for these diseases (Annexure 1). The IDIs were audio-recorded, also after participants consented, notes were taken by a trained note taker. The interviews were conducted face-to-face, and on average, lasted 45 minutes per interview.

### Data analysis

We used Rapid analysis approach (10) to code the data. The Rapid analysis matrix was created in excel sheets, using a deductive approach and the sub-codes and codes were derived first from the data collection tools. Some codes were later inductively added from the data during the coding process. All the coded data were arranged in themes based on the WHO HSS framework. The study team members who collected the data listened to the audio-recorded files and coded the excerpts from the interview in the Rapid analysis matrix. This matrix was later translated from Marathi/ Hindi (local languages) into English. After coding, the data were rearranged using WHO HSS framework where data in each sub-code and later codes was summarized. Data from each code was designated to a WHO HSS framework building block for analysis.

## Results

The qualitative data analysis revealed several barriers and facilitators to the integration of TB, DM, and TC programs in India. These findings provide valuable insights for the program coordinators and implementers in the country. We have presented our results based on the WHO HSS framework’s (2007) building blocks. Each theme is arranged under each of the six building blocks of the framework.

## 1. Service Delivery

Service Delivery entails how healthcare services are delivered and managed. The Indian health system’s current service delivery structure and regulations pose both facilitators and barriers.

### 1.1 Inadequate implementation of protocols

The data confirm the existence of TB-DM and TB-TC frameworks; however, the lack of detailed operational guidelines lead to inadequate collaborative efforts across these programs. While DM screenings are conducted for all TB patients, not all DM patients exhibiting the four-symptom complex (cough of any duration, fever, night sweat, weight loss) undergo screening for TB. This deficiency in service delivery contributes to a neglect of symptoms and a reduce motivation to visit health facilities. Inadequate monitoring, inactive management surveillance, and infrequent public awareness campaigns further hinder to the effectiveness of bi-directional TB-DM screening and smoking control initiatives.

> *“If we see the TB report, 90% of the TB patients are screened for diabetes. There are systems and mechanisms for that. Ni-kshay is there, screening and monitoring is happening meticulously. But if we look at it the other way, there is a gap in the implementation for screening of TB in NCD clinics. Even though the framework is present, there are no operational guidelines as such.” (IDI# 22)*

### 1.2 Lack of Linkage Mechanisms

Barriers for integration of TB-DM-TC program faces barriers due to an inadequate referral system, hindering proper management and continuity of care for patients with co-infections. The absence of communication between primary care facilities and referral hospitals undermines progress and potential enhancements.

> *“…Referral and Feedback mechanisms are not strong at all. Testing is happening in NCDs, but we don’t get any feedback from the TB patients that are sent here in these NCD clinics.” (IDI# 17)* Participants highlighted the absence of links in the referral system, indicating potential disruptions in patient follow-up processes. Issues in rural healthcare delivery encompass non-compliance with the system, self-referrals to doctors, and direct referrals to specialists.

### 1.3 Limited availability of Tobacco Control Centers

The study identified that the integration of TC services into the TB-DM program is hindered by the limited availability of tobacco control centers. Inadequate infrastructure, resources, trained staff, and essential drugs and equipment impeded the uptake and coverage of services. Consequently, there is low awareness and insufficient support for individuals struggling with tobacco addiction.

> *“For tobacco control and counseling, there is no provision for below taluka [block] level, everything happens till block level. Dedicated centers are there along with dentists and counselors, but not all are in a working condition.” (IDI# 9)*

The insufficient presence of trained healthcare professionals further restricted the implementation of the NTCP initiative and contributed to accessibility challenges. Limited awareness regarding the detrimental effects of tobacco has led to a scarcity of Tobacco Control Centers, resulting to a low demand for services and reluctance among individuals to seek access.

### 1.4 Relatively robust program guidelines for TB when compared to DM

All participants unanimously agreed that decentralizing the effective TB control program down to the primary healthcare and private health facility level, serves as an appropriate starting point for the integration of TB-DM-TC programs. The findings indicated that the existing TB program guidelines were firmly established and widely implemented within the Indian health system. These guidelines laid a robust foundation and framework for integrating DM and smoking control services.

> *“NTEP is a well-structured program with good human resources. TB services are provided free of cost. There is a Ni-kshay portal for reporting the patients. The program has strong guidelines and protocols in place. The NCD program is a little weak. Very poor mechanism of referring to the patients.” (IDI# 2)*

The Indian health system lacks well-developed guidelines for DM, with patients managed within general services without a specialized section. This probably leads to abrupt medical attention and assistance to DM patients, creating difficulties in accessing necessary care, even in emergencies.

## 2. Health Workforce

The health workforce theme addresses the availability, responsibilities, training and renumeration for healthcare personnel in integrated care, as well as barriers and facilitators in delivering integrated care.

### 2.1 Well defined responsibilities for implementation of TB program

While the NTEP in India has designated workforce responsible for different levels of implementation, the DM program, conversely, receives minimal attention with no designated staff. The roles and responsibilities within the DM program are unclear. The implementing staff at the public healthcare facilities lack clarity, leading to ambiguity and confusions that hinder effective implementation and the integration of TB and DM screening.

> *“Because of the lack of attention paid to diabetes care, there is no reference guideline. As a result, we have been diagnosing and managing diabetes patients in a haphazard way. Also, the NPCDCS program has many diseases clubbed together because of which there is no separate staff present only for DM.” (IDI# 12)*

### 2.2 Overburdened Human Resources

Healthcare workers in the study areas face overwhelming workloads in implementing routine programs, reporting, procurement, and overseeing critical segments. The current health workforce is insufficient to meet service demands and embrace new initiatives. Introducing integrated TB and DM screening programs might amplify workloads, potentially leading to burnout, fatigue, and stress; such conditions could have a detrimental effect on productivity and efficiency.

> *“There is a lot of workloads on us. We do not even get proper incentives for the amount of work we are doing. We have to keep a check on all the people in our area for all diseases. Apart from our work, sometimes we have to manage PHC work also because there are less people working here.” (IDI# 14)*

### 2.3 Lack of regular training to the Healthcare Personnel

Ensuring regular training for healthcare personnel is crucial for managing TB and DM cases. However, resources limitation and competing priorities often hinder training opportunities, resulting in delayed diagnosis, improper treatment, and poor patient outcomes. The lack of adequate training further contributes to increased turnover rates, intensifying the burden on the healthcare system.

## 3. Medical Products & Technology

The study identified limited issues in the availability and accessibility of essential medicines and diagnostic materials, thereby benefiting the integration of TB, DM, and TC programs.

### 3.1 Supply of medicines & diagnostic kits

A crucial facilitator for the successful implementation of the TB program relies on a smooth supply of medicines and diagnostic kits. The NTEP ensures timely and effective treatment by implementing a drug procurement and distribution system, collaborating with other agencies to ensure quality. *“Yes, sufficient kits are there. For TB, we have a portal which is implemented throughout the country, that is the dispensation module. At what place what drugs are available or are over is known through that portal. Hence immediate actions can be taken.” (IDI# 2)*

Participants acknowledge the current logistic supply chain system as an appealing possibility for long-term medical supply chain administration for both diseases. Contrarily, the frequent stockouts of diabetes management supplies and medicines in India hinder the implementation of diabetes management programs. This leads to interruptions in care, suboptimal control, increased complications, and reduced quality of life. Moreover, these stockouts affect patients’ and providers’ confidence, resulting in a loss of trust in the healthcare system.

> *“For 3 months there has been a shortage of medicines. This happens very frequently when it comes to DM medicines and other supplies. Few months ago, I had no supply of Metformin for 2 months. There is also insufficiency in the glucometer strips due to which the testing is hampered.” (IDI# 15)*

Some respondents express the belief that DM control medicines are prohibitively expensive, resulting in expensive prescriptions from private retail pharmacies. For many economically disadvantaged patients, the inability to afford out of pocket for medicines during stockouts in public health systems leads to lower doses and poor medication adherence. Diagnosing diabetes requires more than merely conducting blood sugar tests; it necessitates medical supplies to ensure effective management in India.

> *“The present integrated medicine supply system, which is managed by the Pharmaceutical Fund and Supply Agency (PFSA), does not adequately accommodate DM supply. We frequently run out of stock. Even at private wholesale providers, the needed commodities for DM care are not always available. Patients are compelled to purchase medications from private retail pharmacies at exorbitant prices.” (IDI#19)*

### 3.2 Ni-kshay Portal, Ni-kshay Poshan Yojana (NPY) and Ni-kshay Aushadhi Yojana (NAY)

The NTEP in India has implemented three initiatives to improve TB programs: Ni-kshay Portal, Ni-kshay Poshan Yojana (NPY), and Ni-kshay Aushadhi Yojana (NAY). Ni-kshay Portal monitors TB patients, providing real-time data on diagnosis, treatment, and outcomes. NPY provides nutritional support and ensures free anti-TB drug supply. These initiatives have not only enhanced the quality of TB care but also facilitated the smooth supply of medicines and diagnostic kits.

## 4. Information

The health information system theme emphasizes the significant of data-driven decision-making and highlights the necessity for a robust health information system. Such a system would play a crucial role in facilitating the integration of TB, DM, and TC programs integration, as well as promoting data sharing across healthcare providers.

### 4.1 Less awareness of HMIS among healthcare professionals and community

The Health Management Information System (HMIS) is a web-based monitoring system in India designed to track health programs, policies, and interventions. It aids a crucial role in grading facilities, identifying aspirational areas, and reviewing Program Implementation Plan (PIP). However, a lack of knowledge about HMIS among healthcare professionals and community members results in its underutilization and hinders data-driven decision-making. Ultimately, this has an impact on healthcare services’ quality and effectiveness.

> *“I have noticed that many healthcare professionals are not familiar with the Health Management Information System and its benefits. This lack of awareness can lead to incomplete and inaccurate data collection, which can ultimately affect the quality of healthcare services. It is important for healthcare professionals to be trained and educated on the HMIS to ensure that accurate data is collected and used to improve the healthcare system.” (IDI# 2)*

### 4.2 Presence of HMIS for every disease

The integration of TB-DM-TC relies on dedicated HMIS systems for each disease. NTEP’s Nikshay tracks TB patients from diagnosis to treatment completion, offering real-time data on cases, treatment outcomes, and drug resistance patterns. NPCDCS’ portal monitors cancer and diabetes patients, while the NTCP’s separate portal contributes to enhance disease control and management across the country.

> *“For NTCP, there is a separate portal where all the data is put till state and district level. Quarterly data is used.” (IDI# 9)*

> *“There is one portal for NCD. The data entry requires around 10 mins for 1 patient. The portal has info about HTN, DM, and about tobacco, and we get to know only if that person is suffering from TB or not.” (IDI# 19)*

### 4.3 Underutilized NPCDCS portal

Our data suggests that the NPCDCS portal is outdated, and underutilized, posing hindrances to integrated programs. With a paper-based reporting system and limited human resources, updating information on a vast array of diseases becomes challenging. This underutilization significantly hinders efficient health management and data recording for DM.

> *“We have different registers for different diseases and we have paper-based records of all the patients when it comes to DM. I do not know about the other two conditions.” (IDI# 14)*

### 4.4 Availability of Resources for IEC activities

Some participants suggested that the ‘Information Education and Communication’ (IEC) activities are crucial for spreading awareness and promoting health-seeking behavior in the community. Resources like funding, trained personnel, and appropriate tools facilitate effective implementation.

*“I believe that availability of resources for IEC activities is crucial for the successful implementation of any health program. These resources can help in creating awareness. This will not only increase the utilization of the system but also improve the quality of data being entered in HMIS.” (IDI# 1)*

## 5. Finance

Financing refers to the mechanisms used to fund healthcare services. The study highlights the need for adequate funding for the integrated program and emphasize the importance of innovative financing mechanisms for sustainability of integrated care.

### 5.1 Availability of Financial Resources

The study highlights the government’s consistent budget allocation for the health sector, ensuring adequate funds for procurement of medicines, equipment, and other resources. This commitment has led to successful implementation of health programs like NTEP and NPCDCS, the recruitment of additional healthcare professionals, and the facilitation of health promotion campaigns and awareness programs, contributing to improved population health outcomes.

> *“For TB funds are sufficient. Occasionally, a delay is observed. For other states, as per disease burden and geographical needs, it is allocated to states/districts. Usually, funds are sufficiently provided to all states.” (IDI# 5)*

### 5.2 Different financial mechanisms for each program

Our data portray different financial mechanisms for each program, potentially hindering the integrated implementation of health programs. These mechanisms contribute to discrepancies in funding allocation, affecting overall health system quality and procurement processes. Additionally, differences in compliance requirements and timelines can result in delays and inefficiencies in implementing the integrated program. To address these challenges, our data suggests the need for an integrated financial system to facilitate holistic resource allocation and management.

> *“I think one of the biggest challenges we face is the separate funding mechanisms for each national health program. It’s difficult to manage the allocation of resources and ensure that each program is adequately funded.” (IDI# 16)*

### 5.3 Limitations in Integrating Programs Based on Financial Schemes

The efficient utilization of resources in the health sector faces a substantial barrier in the form of limitations arising from integrating programs based on financial schemes. The challenge of integrating financial schemes within health programs emerges from variations in funding mechanisms, funds availability, and procedural differences. This disparity can lead in an inequitable distribution of resources, leading to adverse health outcomes and influencing geographical or demographic priorities.

> *“No, funding cannot be integrated. Activities can be integrated together but not funding. There are common cross-cutting strategies or a common budget is provided for few activities. It can be integrated for a particular activity but is not feasible for the whole program.” (IDI# 17)*

### 5.4 Involvement of Private Sector

The involvement of private sectors in health financing can serve as both a facilitator and a barrier to the effective implementation of national health programs, given the diversity opinions of participants. While private sector involvement has potential to enhance quality and coverage, variations across programs lead to discrepancies in funding and resource allocation. Study participants emphasized the importance of managing private sector involvement to complement the goals of national health program.

Participants also emphasized the importance of cross-sectoral planning and involving profit-making firms through Corporate Social Responsibility (CSR) at city district level. This strategy has potential to mobilize resources and funding, ultimately contributing to the improvement of community health.

> *“Yes, it is happening in NTEP. For an increase in private notifications, we are taking help from private agencies, which is currently functional in 35 districts. Current financing is good. Paper based needs are always there. If expenditure is not there only then there is no point. When we get saturated with our own funds then we go to the private sector.” (IDI# 11)*

## 6. Governance & Leadership

Effective integrated care management demands strong governance and leadership, necessitating robust leadership, political commitment and the identification and resolution of barriers and facilitators.

### 6.1 Underdefined systems for involvement of Private Practitioners (PP)

Underdefined systems for private practitioners’ involvement in the integrated health programs pose a hindrance to effective implementation in India, particularly in rural and remote areas. This lack of clarity leads to confusion, uncertainty, and an uneven distribution of healthcare services. The absence of well-defined systems, coupled with a lack of incentives and recognition, hampers the engagement of PP entities in national health programs. Despite functioning independently, the absence of recognition may demotivate their participation. This deficiency can lead to the duplication of efforts and inefficiencies in healthcare service delivery.

> *“PPP is there but has not worked out on all levels. If you want to incorporate private sectors, there are possibilities of PPP like that in TB. It can be done for DM too. There are few NGOs working with NTCP, but there are no private hospitals. That is not the domain for them. While doing PPP, there should be some mandated CSR activities regarding the promotion of harmful effects.” (IDI# 4)*

### 6.2 Well-designed Programs

The effective implementation of healthcare services relies heavily on well-designed national health programs. Programs such as NTEP, NCD and TC programs concentrate on specific health issues, employ evidence-based strategies, and have clear goals, objectives, and structured activities. Consistent monitoring and evaluation mechanism ensure prompt feedback and corrective measures. Study participants unanimously agreed that these programs provide a framework for the coordination and integration of healthcare services, ensuring timely and efficient delivery.

> *“I believe that the current national health programs are well designed and have the potential to make a significant impact on the health of the population. The programs have been developed based on extensive research and consultation with experts in the field.” (IDI# 16)*

### 6.3 History of Tobacco Use Documented in TB as well as DM program

Identifying individuals at risk of developing diseases like TB and DM relies on the crucial assessment of their tobacco use history. These data play a pivotal role in shaping targeted interventions, pinpointing those at risk, and tracking their progress in control and treatment response.

> *“I think documenting the history of tobacco use in TB and DM programs is a really important step in addressing the burden of tobacco-related diseases in India. By identifying patients with a history of tobacco use, we can provide them with targeted interventions and support to quit smoking or chewing tobacco.” (IDI# 9)*

### 6.4 Lack of Optimum Coordination, Communication and Discussion

Our data strongly suggest that the effectiveness of integrated programs rely on collaboration and coordination among national health programs. Facilitated by open communication and discussions, this collaborative approach helps identify areas of overlap, prevents duplication, identifies mutual benefits, and addresses challenges or barriers that could impede the success of the program. Very importantly the cross talk between all three programs for integration was emphasized in the data from both the sites.

While the new integrated program may face challenges in regulation and accountability of policymakers and program managers, strong regulation and collaborative technical standards will likely attract interest from implementers and funding agencies. Most participants also agreed that a TB-DM-TC integrated program within the healthcare delivery system is feasible, provided the DM program’s gap is addressed, and specific tasks are identified for all levels.

> *“I believe that optimum communication and discussion are essential to ensure equal benefit for the participating programs. As a healthcare professional, I have seen first-hand benefits of collaboration and coordination between different programs, especially when it comes to delivering healthcare services in remote and under-served areas. Effective communication ensures that all stakeholders are aware of the programs and their objectives, reducing the duplication of efforts and ensuring that resources are utilized efficiently.” (IDI# 22)*

## Discussions

The primary objective of this study was to investigate potential health system barriers and facilitators that merit consideration in the integration of the TB-DM-TC program. In this section we shed light on the implications of these findings for program coordinators and implementers in India, as well as offer insights into potential strategies for overcoming these challenges and optimizing the integration process.

The absence of collaboration between integrated services for TB-DM and TB-TC is primarily due to the lack of precise operational guidelines, unlike successful integration programs like TB-HIV. To address this issue, leveraging insights from the TB-HIV integration program could facilitate the integration of TB, DM, and TC programs. Success in these programs hinges on their comprehensive, continuous, competent, compassionate, and cost-effective, all planned within the limitations of available resources (11). For illustration, patients with elevated blood sugar levels are typically referred to NCD clinics. However, not all individuals with DM undergo TB testing, as only those with symptoms are screened and directed to TB clinics. This approach may not be comprehensive and could result in missed TB cases in DM patients (12). Considering the high prevalence of DM in the community and the modes of diagnosis and treatment, testing all patients with DM for TB may seem far-fetched. Such a strategy could potentially burden the health system with unnecessary TB testing. To address this challenge, a well-defined approach to TB screening in all DM patients is necessary. This approach should be evidence-based, taking into account the prevalence of TB and DM in the community and the available resources.

The prevailing referral system has proven inadequate in providing feedback on referral outcomes, leading to a lack of communication and follow-up between primary care facilities and referral hospitals. This deficiency indicates a misalignment with current standards, resulting patients identified by the system to fail through the cracks despite considerable efforts. Establishing a stringent system that ensures continuous monitoring of patients registered within the public health framework receive until recovery is crucial. Therefore, establishing efficient communication channels and robust referral systems become imperative. The study also identified that self-referrals to doctors and higher specialization levels hinder healthcare delivery in rural areas. Patients’ non-compliance with the referral system’s hierarchy, coupled with a perceived inadequacy of primary healthcare facilities, leads them to seek care from doctors with higher specialization. This fragmentation of health services can result in worse outcomes. Improving patient education and awareness about the referral system hierarchy is crucial for coordinated care. Strengthening the referral system requires effective communication, coordination, and monitoring mechanisms to ensure compliance with the hierarchy.

Based on its findings, the study highlights that the scarcity of tobacco control centers hinders the integration of smoking control services into the TB-DM program. To improve access, the establishment of more centers, pilot tests and carry out pre-implementation testing in real-world environments are recommended. It is advised to develop context-specific training materials for health personnel to address issues like stigma, ignorance, low knowledge, and illiteracy (13). The Ministry of Health and Family Welfare’s limited tobacco control counseling training initiative has been slow, and further efforts are needed to improve the implementation of NTCP and enhance skills of healthcare professionals. Additionally, the government could explore offering incentives to private healthcare providers for their involvement in tobacco control programs to encourage their active participation.

The study identified potential opportunities within the Indian health system to implement the proposed TB-DM-TC strategy, which is in accordance with prior studies. Notably, the well-organized TB control programme (14) and the partially institutionalized TB-DM screening (8,15) serve as foundations for potential integration. This finding underscores the existence of a robust framework upon which to the integration of DM and tobacco control programs. The well-established and widely in the Indian health system provides an opportunity to leverage existing infrastructure and expand the scope of care to include DM and TC services.

Despite established regulations and standards, service delivery for TB/DM comorbidity remains deficient. This is a matter that cannot be neglected, particularly as India strives to achieve TB-free status five years ahead of global targets. Vigilance within healthcare systems is paramount, and the community must be informed about this critical issue.

In line with them, the study identifies the main barriers to integrated care: inadequate knowledge of healthcare providers (1,7,8); lack of diagnostic tools and equipment (especially DM kit and medicine supplies); and insufficient attention to DM control and smoking control program. The lack of knowledge in HMIS contributes to these barriers, impacting the motivation and engagement of healthcare workers and community members in utilizing the system. Healthcare workers may perceive HMIS as an additional burden to their already demanding workload, leading to resistance in its implementation. Therefore, effective communication strategies are critically needed. Moreover, the community’s underappreciation of TB/DM comorbidity leads to a lack of awareness and knowledge about tobacco and alcohol use. Addressing this issue requires community-based interventions that aim to create awareness, reduce stigma, and motivate individuals to seek healthcare services. Additionally, health workers should be educated on the importance of intervening in smoking for TB patients, and providers must receive proper training (13).

The study also highlights the importance of cross-program and cross-sectoral collaboration for efficient execution and sustainability in integrated programs. This collaborative approach not only promotes resource efficiency but also minimizes duplication and ensures the continuity of care. The significance of multisectoral coordination and cooperation, particularly involving private primary care providers in the collaborative management of TB-DM, has been discussed in Indonesia-based study (16) and consistently advocated by other studies (1,17).

The establishment of culture-sensitive infrastructure is crucial for the success of preventive operations, such as screening, to ensure user engagement and early acceptance. The suggestion for tri-directional screening of TB-DM-smoking at community outreach and primary healthcare facilities, along with intensive health promotion activities to increase community health literacy (7). However, if the Information Education and Communication (IEC) messages are not culturally appropriate or not translated into local languages, they may not effectively reach the target population. Hence, modifying or adapting the IEC materials to the local context is necessary. Additionally, enhancing the national health information system is recommended to address reporting and surveillance issues (1,17). Implementers and decision-makers should collaborate to create an integrated health information system, establish shared objectives, and provide training for care providers on reporting and recording. This study offers decision-makers implementation tactics for tailored interventions within the resource-constrained Indian health system.

## Conclusion

Overall, this study outlines the barriers and facilitators to implementing an integrated model. This study has illuminated the substantial challenges in integrating the TB, DM, and TC programs within India’s public health systems. The findings underscore the urgent need for a comprehensive and cohesive approach to tackle India’s burden of TB, DM and tobacco smoking. Furthermore, it highlights the importance of establishing operational guidelines to foster collaboration among various public health initiatives, setting a precedent for integrating additional programs within India.

Our study suggests leveraging the established NTEP infrastructure as a robust foundation for incorporating DM and TB services within primary care, underscoring the necessity of addressing deficiencies in service delivery for TB/DM comorbidity, especially in light of India’s aspirations to achieve TB-free status. Also, it strongly advocates for the convergence of the three programs into a single-window approach, along with the establishment of effective feedback and referral systems, and the development of harmonized digital data entry platforms. A multifaceted approach is recommended for policymakers and management of the healthcare system use.

To enhance user engagement and facilitate early detection, the study proposes the implementation of culturally sensitive infrastructure, along with community-based interventions and health promotion activities. Moreover, it highlights the significance of enhancing the national health information system and implementing tailored interventions within resource-constrained environments for the successful integration efforts. Additionally, the study emphasizes the necessity of ongoing research to continually evaluate the efficacy of integrated models in addressing complex health issues in the Indian context.

### Recommendations

Participants recommended establishing an independent integrated TB-DM-TC program using existing resources, developing joint technical guidelines for standardization, financial planning, and national budgeting. The study participants also advocated for the integration of patient record software onto a single platform with a unique identification number. Furthermore, availability of single visits for TB patients with comorbidities, enabling crosstalk of all three programs, involving private practitioners in screening and sensitizing them for Ni-kshay registration should be done. There must be rigorous implementation of the feedback and referral system, training modules incorporating joint planning, implementation, monitoring, and evaluation. Also, there should be provisional psychosocial support for patients, and enhance the use of digital health technologies among communities. These recommendations aim to improve patient outcomes and efficiency in the national health programs.

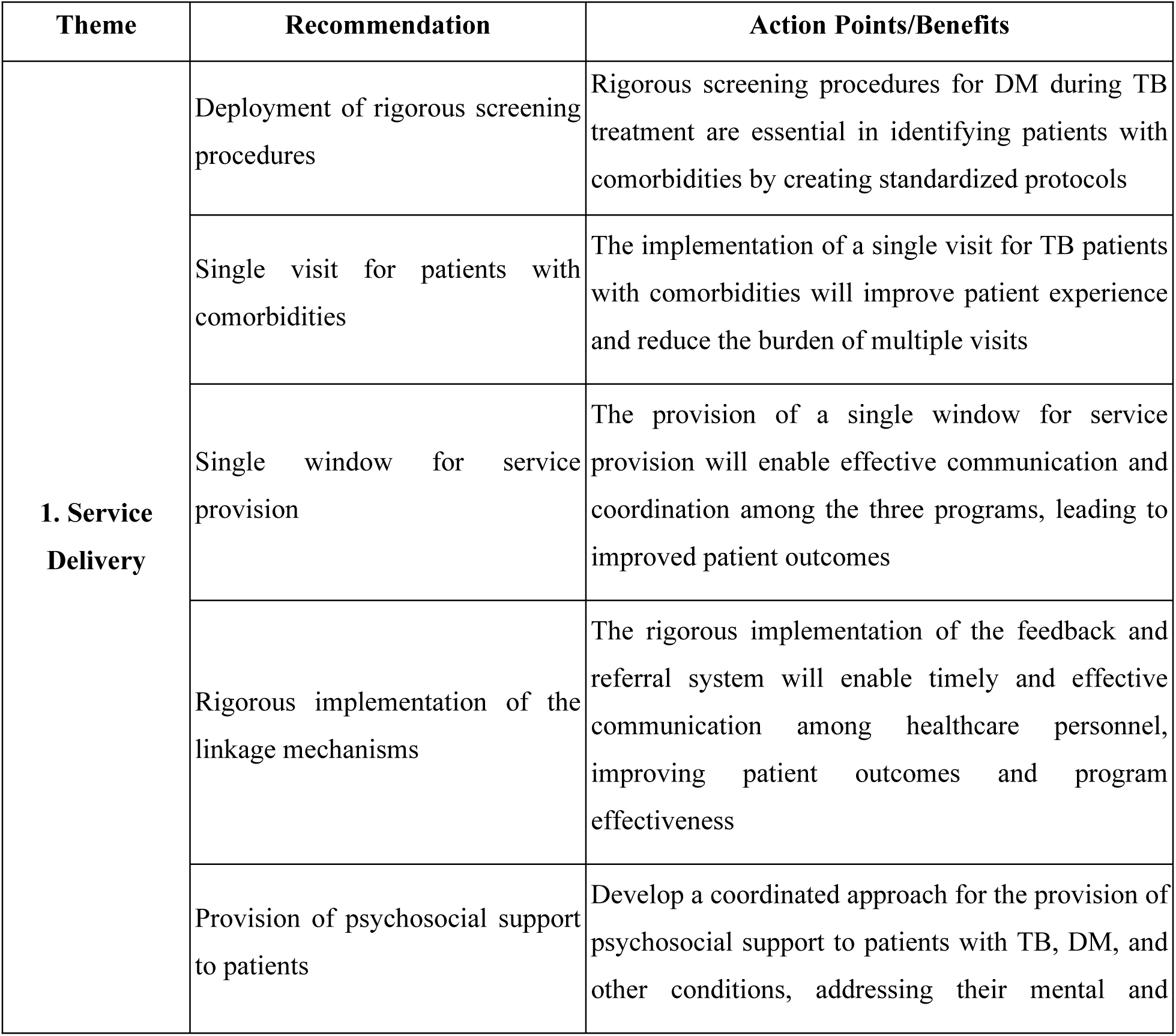
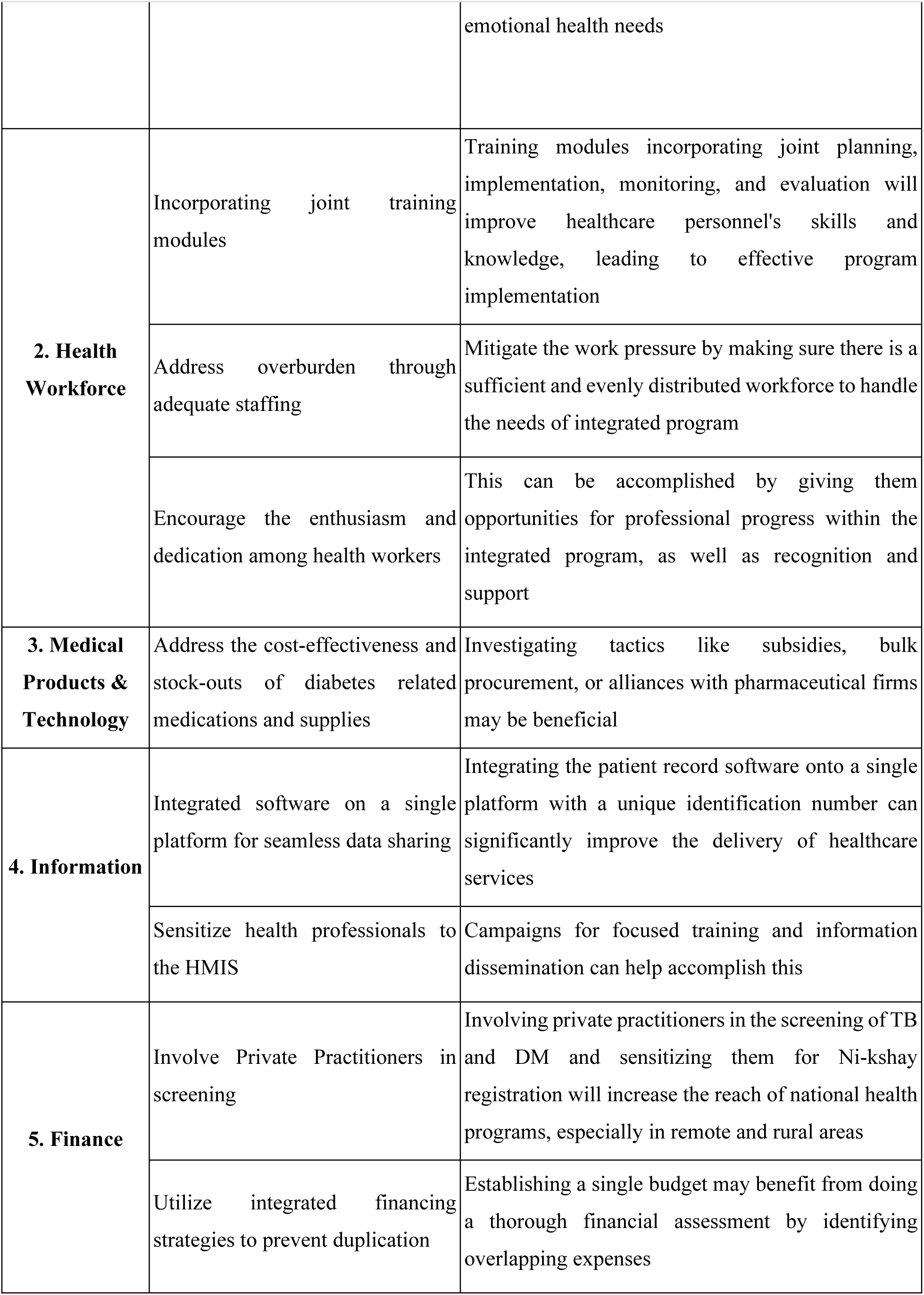
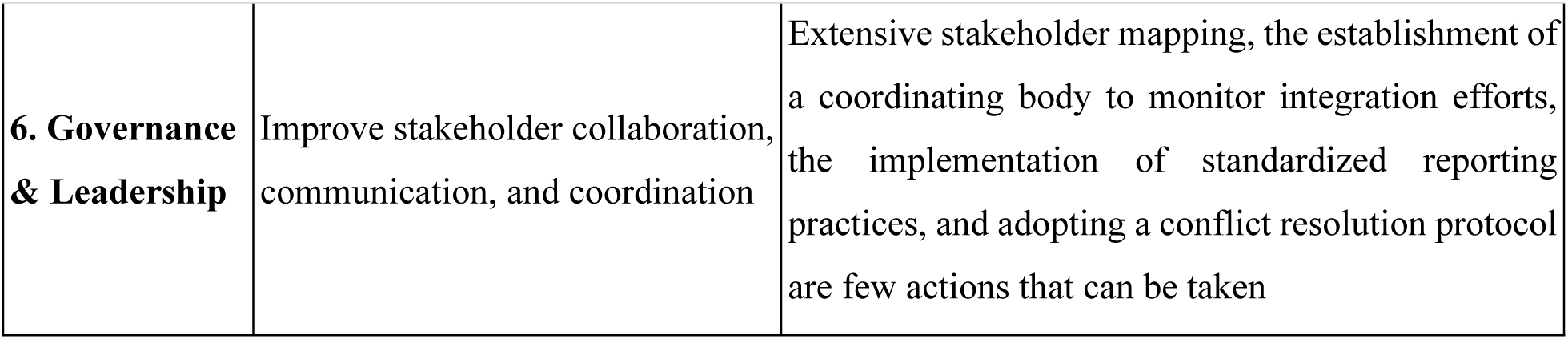

## Data Availability

All the data underlying our findings are included within the manuscript and the coded data will be made available on request.

## Annexure-1

Following is the study tool used for collecting the data:

### Key-Informant Interview

#### Objectives

To understand the Existing programs in relation to TB, DM, and smoking control programmes
To explore the facilitators and barriers for the integration of the three programmes at the national, provincial/state and district levels

#### Framework

The key information interview is developed based on the six building blocks of the health system as the main framework, which includes the following dimensions: (1) Service delivery, [2) Health workforce; (3) Health information system: (4) Financing; (5) Governance and leadership; and (6) Access to essential medicine.

### Interview Guide

#### Opening questions

1. Which institution are you working at?
2. What is your current position and responsibility?
3. How are the states (province/district/city) dealing with the dual burden of communicable and non-communicable diseases?
4. Has the PIP (Programme Implementation Plan) been Tailored to the disease burden in the state and district level?
5. Is there a budget allocation?
6. Are you doing any implementation research in this regard?

Please indicate how often have your institution been working together with the folIowing institutions.

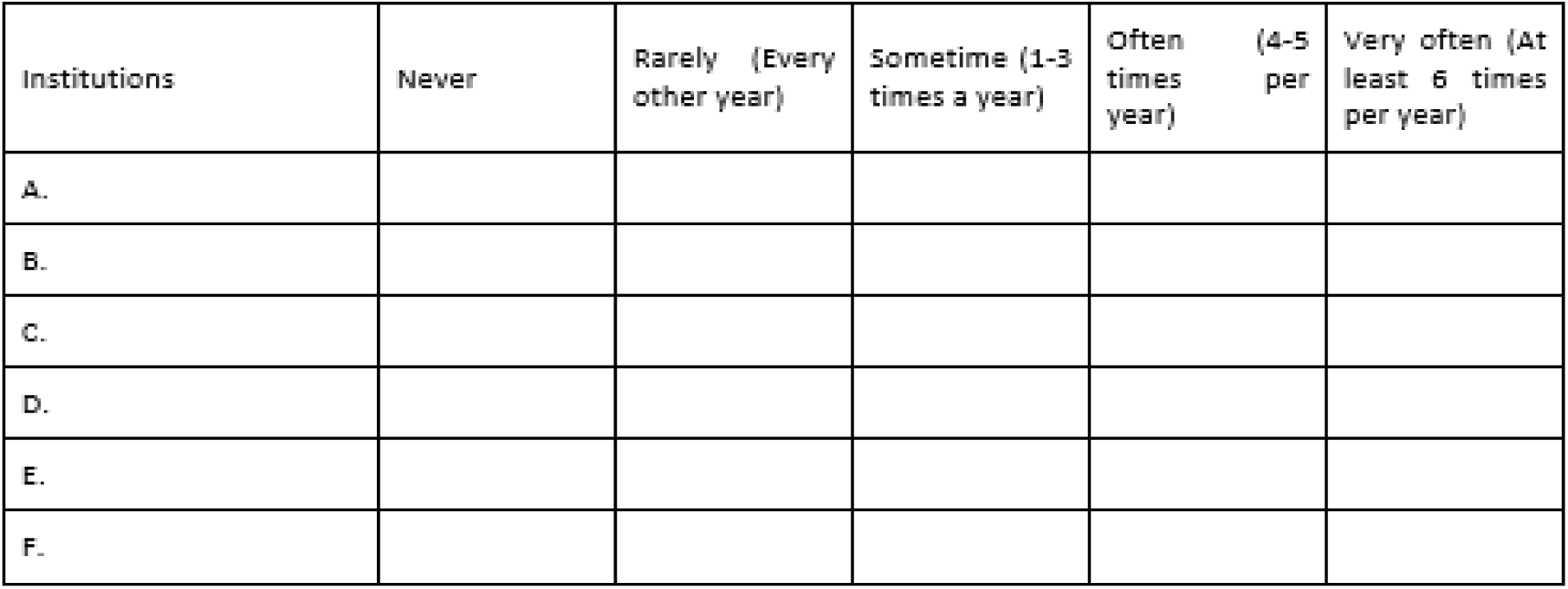

#### Interview questions related to the health system block

#### Service delivery

◦ How is the bi-directional screening of TB and DM organized at different levels of the health system? Is it working well? Does it require more strengthening? What do you think will help enhance integration?
◦ What are the opportunities that you see and the barriers they face when they plan to do the TB and DM screening?
◦ How do we ensure the comprehensive screening of TB and DM through both active an d passive screening?
◦ What is the current situation regarding the above and what are suggestions for more comprehensive integration of communicable and non-communicable disease programme?

#### Health workforce

◦ The human resources: Do they have the personnel to do the screening/disease management? Sufficiency? Task shifting?
◦ Training: do you have the specific knowledge to do the integrated disease control program (eg. TB-DM)? Have you been trained in conducting scree ring 3rd disease control?
◦ Coordination between the different disease program staff

#### Health information system

◦ Current disease program information system: the linkages of infrastructure in the different programme, the load of recording and reporting
◦ The current plan of an integrated information system
◦ Are the data used in the coordination meeting?
◦ Are they using the data for evidence-based decision making?

#### Financing

◦ Fund-flow mechanisms - do the fund come to the district/PHC level?
◦ Are government funding available for TB/DM/Tobacco control programme? Assess differences between the states/districts
◦ Wcu d you think the integration of Communicable anc Non-Communicable program is tea sib e based on the financing scheme? Why?
◦ Is the private sector involved in financing the integrated programs? Continuity of the funding?

#### Governance and Leadership

◦ What kind of inter-departmental and inter-sectoral cooperations/coalitions they have within and outside the government?
◦ Are there guidelines? Have they been disseminated? What do you think about the guideline?
◦ Leadership and coordination for sharing resources (diagonal sharing) - whether the rich resources would like co-share with another program
◦ Fund flow mechanism
◦ What are the common challenges you would face in the integration of the programme?
◦ Is there any policy to involve private sectors and NGOs in the integration of communicable and non-communicable diseases control programme.

#### Access to essential medicine

◦ Do you have sufficient screening and diagnosis kits (sputum test, GeneXpert, et) for tuberculosis and diabetes?
◦ Do you have sufficient essential medicine for treatment for tuberculosis and diabetes?
◦ What are the estimated costs of treating a patient with TB and/or DM?

Please think or all the actions that **your institution** can do to facilitate the integration or TB, DM and tobacco programme. There is no right and wrong artswer, please list all the actions that you can think of.

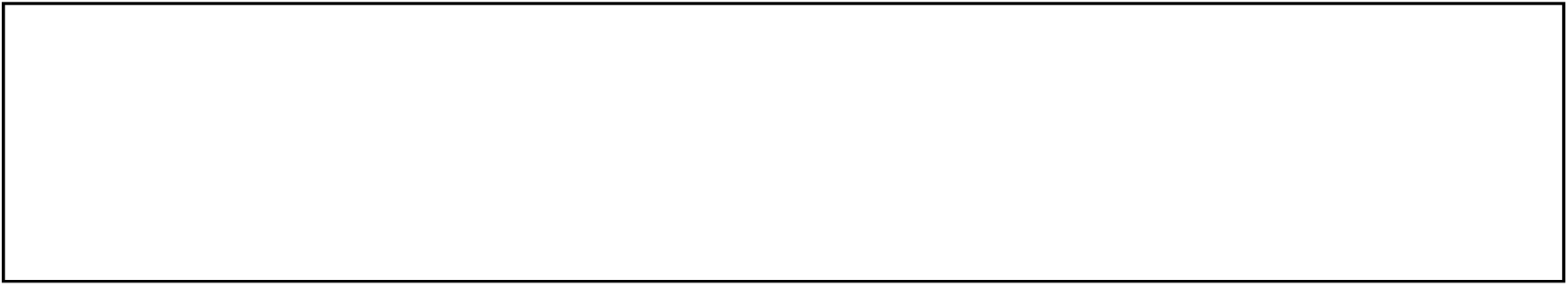

Please think of all the actions that other institutions mentioned above can do to facilitate the integration of TB, DM and tobacco programme. There is nc right and wrong answer, please list a I the actions that you can think of.

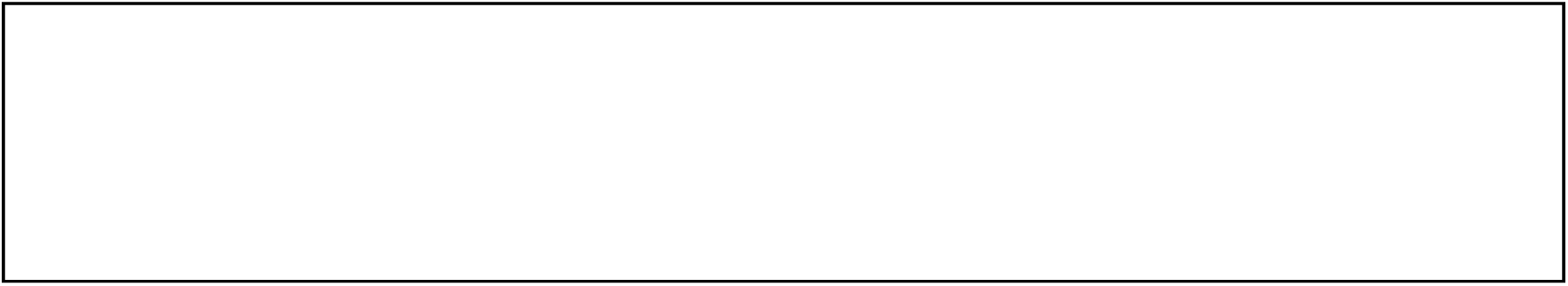

